# Meningovascular inflammation in cerebral amyloid angiopathy-related cortical superficial siderosis

**DOI:** 10.1101/2025.09.24.25336546

**Authors:** Philip Arndt, Eya Khadraoui, Sebastian Müller, Katja Neumann, Hendrik Mattern, Sven G. Meuth, Valentina Perosa, Andreas Charidimou, Stefanie Schreiber

## Abstract

The role of inflammation in cortical superficial siderosis (cSS), a marker of cerebral amyloid angiopathy (CAA) linked to high hemorrhage risk, is unclear. We examined 14 patients with cSS using 3T post-contrast vessel wall MRI (VWI) and CSF analysis. Although only 31% met current CAA-ri criteria, 86% showed vessel wall enhancement or sulcal hyperintensities near cSS, frequently extending beyond. Four patients with follow-up VWI demonstrated corticosteroid-responsive regression of inflammation. CSF albumin quotients, indicating blood–brain barrier dysfunction, correlated with MRI inflammation scores. These findings reveal subclinical meningovascular inflammation in cSS and support VWI for detecting a broader CAA-related inflammation spectrum.

## Background

Sporadic cerebral amyloid angiopathy (CAA) is typically an age-related, slowly progressive, and largerly non-inflammatory small vessel disease, characterized by the deposition of beta-amyloid (Aβ) within the walls of cortical and leptomeningeal vessels.^1^ CAA is now well established as a common cause of lobar intracerebral hemorrhage (ICH) and a major contributor to cognitive impairment in older individuals. In a subset of patients, however, preexisting CAA may rarely trigger an autoinflammatory response directed against vascular Aβ, resulting in inflammatory variants of the disease.^2^ This condition, known as CAA-related inflammation (CAA-ri) in its typical form, presents with a distinct clinical and radiological phenotype, often marked by acute or subacute neurological symptoms and imaging findings that diverge from the typical profile of sporadic CAA. According to current diagnostic criteria, a diagnosis of CAA-ri requires progressive neurological symptoms accompanied by confluent, often asymmetric, white matter hyperintensities extending into the subcortical white matter on MRI.^3^

However, recently the concept of CAA-ri spectrum disorders was introduced to acknowledge a broader continuum of neuroinflammatory involvement in sporadic CAA.^2^ This emerging framework suggests that neuroinflammation may play an underrecognized role in cases of seemingly sporadic (i.e. non-inflammatory) CAA and offers a more inclusive toolkit to diagnose CAA-ri spectrum disorders. The concept was grounded on a retrospective case series of six patients with cortical superficial siderosis (cSS) and a clinical presentation of prolonged, unremitting CAA-related transient focal neurological episodes (TFNEs), as well as other cases increasingly encountered in CAA clinical practice, who do not meet the current diagnostic criteria for the typical CAA-ri syndrome.^4, 5^ While these patients failed to fulfill established, but arguably limited in scope, CAA-ri diagnostic criteria, they nonetheless exhibited inflammatory features on imaging - including sulcal, leptomeningeal, cortical vessel wall, and parenchymal enhancement on contrast-enhanced and vessel wall MRI - suggestive of underlying blood– brain barrier (BBB) dysfunction and neuroinflammation.^4^

We present a well characterized series of 14 patients with CAA-related cSS, encompassing a range of CAA-related clinical phenotypes, and available cerebrospinal fluid (CSF) data, who underwent post-contrast MRI with high-resolution vessel wall imaging (VWI). This series provides further evidence for a cSS-linked, widespread neuroinflammatory process across diverse clinical presentations, lending additional support and validity to the emerging concept of CAA-ri spectrum disorders.

## Methods

### Patients

This is a retrospective consecutive case series of 14 patients presenting with CAA-related cSS and various CAA-related clinical symptoms who underwent VWI and post-contrast MRI at the Department of Neurology, Otto-von-Guericke University in Magdeburg between May 2021 and June 2025. The local ethic committee (No.331 07/2017, addendum 11/2021) approved this study.

### MRI data

Patients underwent a standardized MRI (median 5 days after clinical presentation, range: 0–8 days), including T1-weighted, T2-weighted, FLAIR, T2^*^-weighted sequences (gradient-recalled echo), diffusion-weighted imaging (DWI), and pre- and post-contrast VWI (black blood imaging). High-resolution VWI was acquired after administration of Gadovist using a transversal, single-slab, non-selective 3D turbo spin echo sequence with generalized autocalibration partial parallel acquisition for image acceleration and spectral attenuated inversion recovery for fat suppression. Whole-brain coverage was achieved with a 200 mm field of view, isotropic voxel size of 0.8 mm, repetition time of 700 ms, and echo time of 23 ms. All scans were performed on a 3-Tesla MRI system (Philips Achieva, Best, Netherlands).

Two senior neuroradiologists independently analyzed all sequences. The Auriel criteria were applied for the assessment of CAA-ri^3^. Vascular lesions were classified according to the STRIVE-2 criteria.^6^ The Fazekas score was used to quantify white matter hyperintensities (WMH).^7^ The global cortical atrophy scale to assess atrophy,^8^ and the Microbleed Anatomical Rating Scale to evaluate cerebral microbleeds (CMB).^9^ VWI was assessed separately for large vessels (A1–A3, M1–M3, P1–P3, vertebral arteries, basilar artery), medium-sized leptomeningeal vessels, and small cortical vessels. The number of contrast-enhancing vessel walls was counted. All enhancement findings were cross-checked with pre-contrast VWI sequences to minimize artifacts. If uncertainty remained regarding whether an enhancing structure was arterial, it was not classified as vessel wall enhancement. Veins were differentiated based on their typical anatomical trajectory, continuity, and morphology. Sulcal hyperintensities or inflammatory parenchymal edema was evaluated on native FLAIR images. Since sulcal hyperintensities may also reflect (sub)acute convex subarachnoid hemorrhage, corresponding signal abnormalities were carefully checked on native T1-weighted images and excluded in all patients.

To generate a composite score for inflammation, we propose to sum the following results: leptomeningeal, and cortical vessel wall enhancement on VWI, and sulcal hyperintensities and inflammatory parenchymal edema on native FLAIR (for each feature: presence - 1 point; no - 0 points). This resulted in a composite MRI inflammation score ranging from 0 to 4.

### Cerebrospinal fluid

Lumbar puncture was performed as part of the diagnostic workup. CSF samples were centrifuged at 4°C, aliquoted, and stored at –80°C. Biomarkers including Aβ_42/40_ ratio, phosphorylated Tau 181 (pTau), total Tau (tTau), and neurofilament light (NfL) were measured using automated immunoassays (LUMIPULSE® G600 II, Fujirebio). Pathological thresholds were Aβ_42/40_ < 0.69, pTau > 70 pg/mL, and tTau > 404 pg/mL.^10^ CSF and serum albumin and IgG were analyzed by rate nephelometry (Immage 800, Beckman Coulter). The albumin quotient (×10^−3^; Qalb) assessed BBB dysfunction, and the IgG index evaluated intrathecal IgG synthesis.^11^

### Statistical analysis

Continuous variables are presented as median (range), and categorical variables as proportions. CSF markers of BBB dysfunction (total protein, Qalb) were correlated with the composite MRI inflammation score using linear regression. Analyses were performed with GraphPad Prism 10.0.3.

#### Results

Demographic, clinical and imaging characteristics of the included patients are summarized in **Table 1**. The median age was 80 years (range: 66-91 years), and seven patients (50%) were female. All patients exhibited cSS, with 71% showing a disseminated pattern (≥4 sulci affected) and 29% a focal pattern (≤3 sulci). Four patients met the current diagnostic criteria for CAA-ri.^3^ Of note, 12 (86%) patients fulfilled the recently suggested criteria for CAA-ri spectrum disorders. Clinical presentations included (sub)acute focal neurological deficits due to ICH, SAH, or vasogenic edema, progressive cognitive decline, TFNE, in one patient transient visual symptoms that justified MRI, and in one patient a focal neurological deficit without an acute lesion (**Table 1**).

**Table 1.** Characteristic of patients with cortical superficial siderosis who meet vs. do not meet current criteria for CAA-related inflammation.

|  | All patients<br>n = 14 | Not meeting clinical and<br>imaging CAA-ri criteria<br>n = 10 | Meeting the clinical and<br>imaging CAA-ri criteria<br>n = 4 |
| --- | --- | --- | --- |
| Age | 80 (66 - 91) | 82 (66 - 91) | 78 (69 - 88) |
| Female | 7 (50%) | 6 (60%) | 1 (25%) |
| **Clinical presentation** |  |  |  |
| Progressive cognitive decline | 3 (21%) | 3 (30%) | 0 (0%) |
| Lobar ICH | 3 (21%) | 3 (30%) | 0 (0%) |
| Focal motor deficit with SAH | 2 (14%) | 2 (20%) | 0 (0%) |
| TFNE | 2 (14%) | 1 (10%) | 1 (25%) |
| Focal neurological deficit with<br>vasogenic edema | 2 (14%) | 0 (0%) | 2 (50%) |
| Transient visual symptoms | 1 (7%) | 0 (0%) | 1 (25%) |
| Focal neurological deficit without<br>acute lesion | 1 (7%) | 1 (10%) | 0 (0%) |
| **CAA markers on MRI** |  |  |  |
| Disseminated cSS | 10 (71%) | 8 (80%) | 2 (50%) |
| Number of lobar CMB | 5 (0-400) | 7 (0-23) | 10 (0-400) |
| Severe CSO PVS (>20) | 14 (100%) | 10 (100%) | 4 (100%) |
| Multispot WMH pattern | 12 (86%) | 9 (90%) | 3 (75%) |
| **Other CSVD markers on MRI** |  |  |  |
| WMH Fazeka scale [0-6] | 5 (2 - 6) | 5 (3-6) | 4.5 (2-6) |
| Severe BG PVS (>20) | 7 (50%) | 5 (50%) | 2 (66%) |
| Presence of lacune | 9 (64%) | 6 (60%) | 3 (75%) |
| Global cortical atrophy score | 2 (1-3) | 2 (1-2) | 2 (1-3) |
| **Cerebrospinal fluid** | n = 10 | n = 7 | n = 3<sup>a</sup> |
| A+T-N- | 4 (40%) | 3 (43%) | 1 (33%) |
| A+T-N+ | 4 (40%) | 3 (43%) | 1 (33%) |
| A+T+N+ | 2 (20%) | 1 (14%) | 1 (33%) |
| Neurofilament light [pg/mL] | 1866 (895-13619) | 1602 (895-7919) | 2968 (2017-13619) |
| Protein [g/L] | 0.52 (0.37-1.31) | 0.51 (0.37-1.31) | 0.50 (0.49-0.52) |
| Albumin quotient | 10.9 (5.3-14.0) | 9.64 (5.3-14.0) | 10.9 (8.3-13.0) |
| IgG-index | 0.50 (0.45-0.70) | 0.50 (0.45-0.60) | 0.70 (0.50-0.70) |

Data are presented as median (range) or as proportions.
<sup>a</sup>One case underwent lumbar puncture and CSF analysis after anti-inflammatory treatment (protein 0.49 g/L, albumin quotient 8.3, IgG-index 0.70).
Abbreviations: BG, basal ganglia; CAA-ri, cerebral amyloid angiopathy related inflammation; CMB, cerebral microbleed; CSO, centrum semiovale; cSS, cortical superficial siderosis; CSVD, cerebral small vessel disease; DWI, diffusion weighted imaging; ICH, intracerebral hemorrhage; MRI, magnetic resonance imaging; PVS, perivascular spaces; TFNE, transient focal neurological episodes; SAH, subarachnoid hemorrhage; WMH, white matter hyperintensities.

The majority of patients showed neuroimaging evidence of inflammation on VWI, including leptomeningeal enhancement (n=12 patients, 86%) or cortical vessel wall enhancement (n=11 patients, 79%), while inflammatory parenchymal (vasogenic) edema was less frequent (n=3 patients, 21%) (**Table 2**). Among those with vessel wall enhancement, sulcal hyperintensities were visible on native FLAIR in all 12 patients, without corresponding hemorrhage on T1-weighted MRI. Both vessel wall and sulcal enhancement were pronounced in areas affected by cortical superficial siderosis (cSS), although vessel wall enhancement was also observed distant from cSS (**Figure 1-2**). Notably, nine patients exhibited diffuse vessel wall enhancement throughout the brain. Enhancement of large-caliber arteries was seen in six patients (43%).

**Table 2.** MRI markers of inflammation.

| Case | Cortical superficial siderosis | Parenchymal (vasogenic) edema | Sulcal HI on native FLAIR | LME | Counts of LM VWE | Counts of cortical VWE | Large VWE | Treatment for inflammation |
| --- | --- | --- | --- | --- | --- | --- | --- | --- |
| *Cases not meeting the CAA-ri criteria* |  |  |  |  |  |  |  |  |
| 1 | bilateral, disseminated | no | yes | bilateral, diffuse | >100 | 4 | right M3 | corticosteroids for 2d |
| 2 | right, disseminated | no | yes | bilateral, diffuse | >100 | 8 | no | no |
| 3 | bilateral, disseminated | no | yes | bilateral, diffuse | >100 | 3 | no | corticosteroids for 3d |
| 4 | bilateral, disseminated | no | yes | bilateral, diffuse | 51 | 6 | right M3 | no |
| 5 | bilateral, disseminated | no | yes | bilateral, parieto-temporal | 41 | 0 | no | corticosteroids for 3d |
| 6 | bilateral, disseminated | no | yes | bilateral, diffuse | 26 | 2 | no | no |
| 7 | bilateral, disseminated | no | yes | bilateral, diffuse | 28 | 3 | Bilateral M2 and M3 | corticosteroids for 3d |
| 8 | bilateral, disseminated | left, parietal | yes | bilateral, fronto-parietal | 16 | 5 | left M1, right M2 | corticosteroids for 3d |
| 9 | left, focal | no | no | No | 0 | 0 | no | corticosteroids for 3d |
| 10 | right, disseminated | no | no | No | 0 | 0 | no | corticosteroids for 3d |
| *Cases meeting the CAA-ri criteria* |  |  |  |  |  |  |  |  |
| 11 | left, focal | left, parietal | yes | bilateral, diffuse | >100 | 3 | right M1 & A3, left M2 | corticosteroids for 3d |
| 12 | left, focal | no | yes | bilateral, diffuse | 71 | 7 | no | corticosteroids for 5d |
| 13 | bilateral, disseminated | no | yes | bilateral, diffuse | 39 | 3 | bilateral M2 | No |
| 14 | right, disseminated | right, frontal | yes | right, fronto-temporal | 27 | 2 | no | corticosteroids for 5d |

Abbreviations: CAA-ri, cerebral amyloid angiopathy related inflammation; d, days; HI, hyperintensities; LME, leptomeningeal enhancement, MRI, magnetic resonance imaging; VWE, vessel wall enhancement

**Figure 1.**
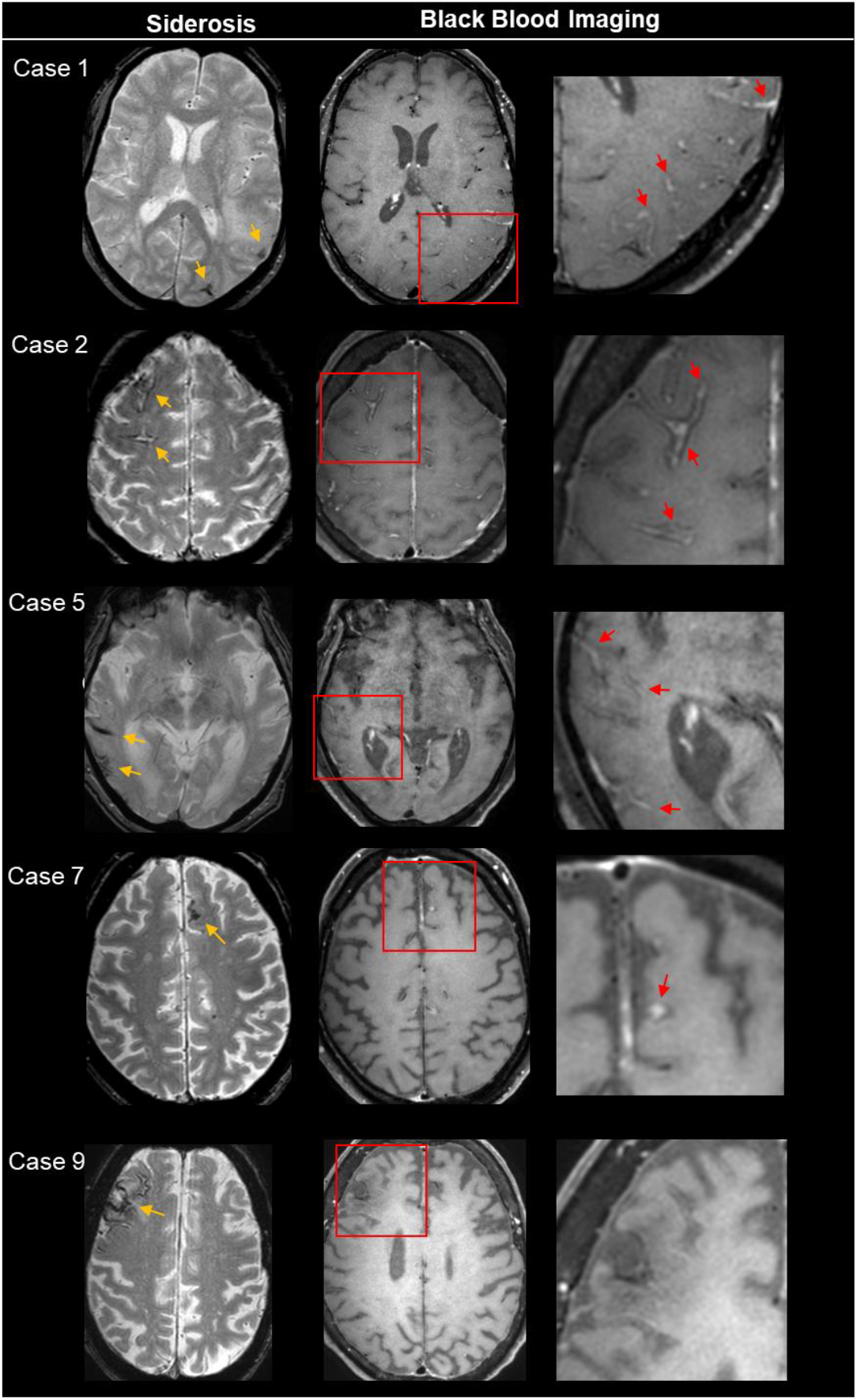
Anatomical overlap of cortical superficial siderosis and vessel wall enhancement on brain MRI. Five representative cases illustrate the variability in the severity of vessel wall enhancement on post-contrast vessel wall imaging. Cases 1 and 2 demonstrate severe enhancement with typical “donut-signs”, cases 5 and 7 show moderate enhancement, and case 9 shows no detectable vessel wall enhancement. Corresponding clinical and imaging variables for each patient are demonstrated in **Table 1-2**. Image interpretation was guided by pre-contrast sequences and anatomical landmarks to avoid venous misclassification.

**Figure 2.**
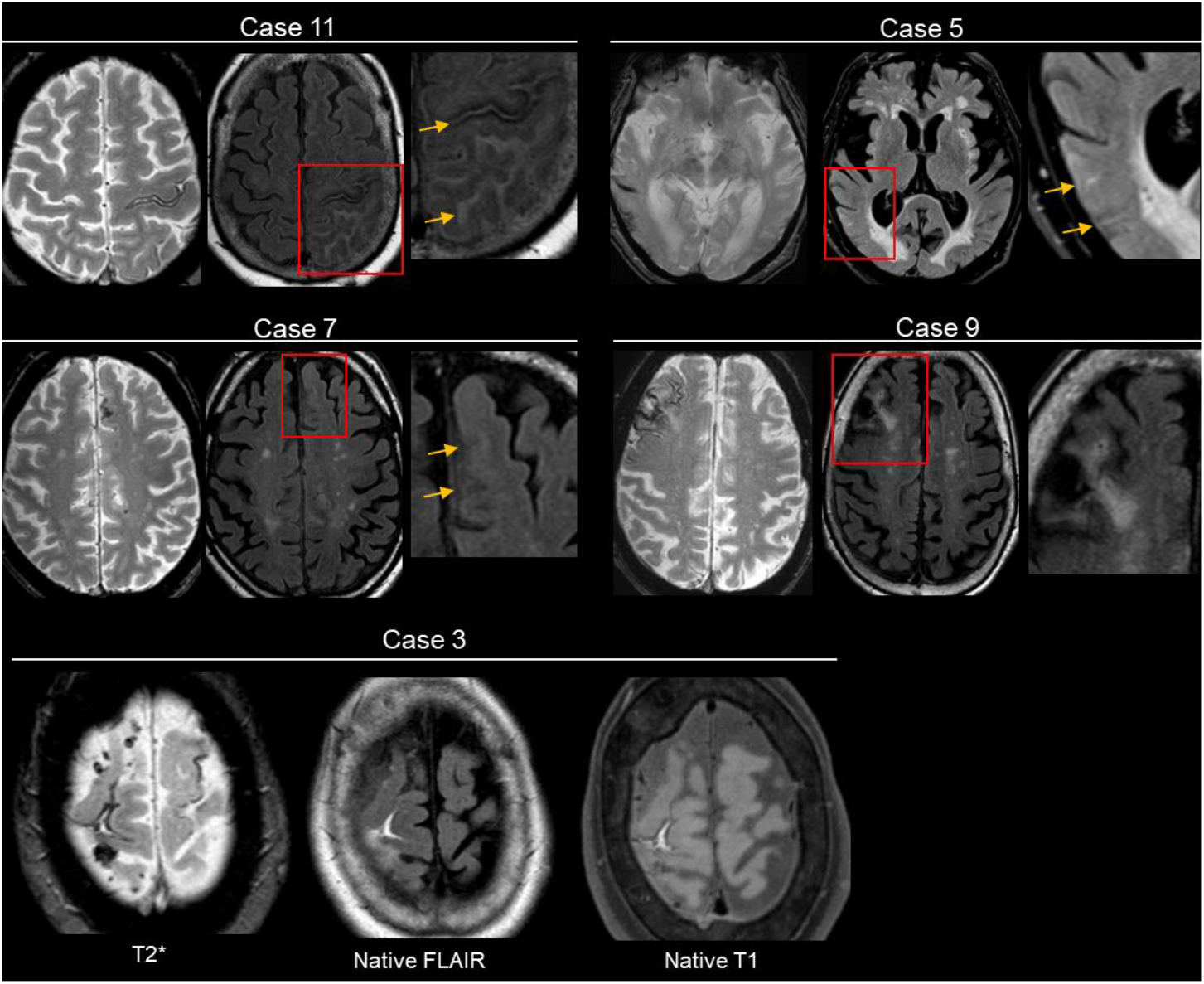
Anatomical overlap of cortical superficial siderosis and sulcal hyperintensities on native FLAIR. Five representative cases illustrate the variability in sulcal hyperintensities extend on native FLAIR sequences. Case 11 shows a severely affected patient meeting current clinical and radiological CAA-ri criteria. Cases 5 and 7 display localized sulcal hyperintensities that correspond to the side of siderosis, while case 9 shows no detectable sulcal hyperintensities. Of note, sulcal hyperintensities may also reflect (sub)acute subarachnoid hemorrhage, as in case 3, where corresponding signal abnormalities are also visible on native T1-weighted sequences. Clinical and imaging variables for each case are summarized in **Table 1-2**.

In four patients treated with high-dose corticosteroids for 2 to 5 days, follow-up MRI demonstrated a reduction in vessel wall enhancement and parenchymal edema. Interestingly, two cases (one with focal, one with disseminated cSS; both not meeting the current CAA-ri criteria; case 8 & 9) did not show an inflammatory process in the initial MRI on VWI or native FLAIR MRI.

CSF samples were available for ten patients, nine of whom obtained prior to initiation of anti-inflammatory treatment. All patients were within the Alzheimer’s disease continuum (A+T-N-n=4, A+T-N+ n=4, A+T+N+ n=2). Elevated total protein levels and increased Qalb values were consistent with BBB dysfunction **(Table 1)**. Notably, Qalb demonstrated a positive association with the MRI-based inflammation score (R^2^ = 0.54, p = 0.024).

## Discussion

This case series provides further supports to the concept of CAA-ri spectrum disorders,^2^ highlighting the presence of an inflammatory process with associated BBB dysfunction in patients with CAA-related cSS, regardless of clinical phenotype. Using high-resolution VWI and native FLAIR MRI, we identified vessel wall enhancement in close anatomical proximity to cSS in most patients, suggesting a spatial and potentially pathophysiological link between cSS and underlying meningovascular inflammation. Notably, we observed substantial variability in the extent and severity of inflammatory changes across patients, including both those who did and did not fulfill current diagnostic CAA-ri criteria. Two patients with cSS lacked neuroimaging evidence of inflammation entirely, underscoring the heterogeneity of presentations. Still, our findings provide a pragmatic imaging-based approach to recognizing neuroinflammation in CAA, particularly in cases with cSS, and lay the groundwork for more refined pathophysiological models of CAA-ri spectrum disorders. Ultimately, this framework may help guide the selective use of immunomodulatory therapy aimed at reducing CAA-related vascular injury and bleeding, an unmet need in the field.

Importantly, ten of fourteen patients in our series did not meet the current CAA-ri diagnostic criteria, which remain anchored solely on the presence of confluent, asymmetric WMH on FLAIR MRI sequences.^3^ Yet, eight of these ten cases (80%) showed clear imaging evidence of meningovascular inflammation—an observation consistent with prior findings in a cSS-TFNE case series, where five of six patients (83%) demonstrated inflammation on VWI despite not meeting formal CAA-ri criteria.^4^ These findings suggest that contrast-based VWI offers sensitivity for detecting inflammation in CAA and supports the notion of their inclusion in the recently suggested framework of CAA-ri spectrum disorders.^2^ The revised framework must also better reflect the diversity of neuroinflammatory patterns across CAA presentations - particularly in patients with cSS, who may harbor clinically significant, yet under-recognized, inflammation. In parallel, the integration of fluid biomarkers reflecting meningovascular inflammation - including indicators of endothelial activation, BBB dysfunction, or specific inflammatory signaling pathways - could enhance diagnostic precision and complement imaging-based assessments. Together, these tools may help redefine the boundaries of CAA-ri spectrum disorders, moving beyond the classical syndrome to recognize atypical, yet treatable, presentations. From a pathophysiological standpoint, cSS in CAA is thought to result from recurrent, self-limiting leptomeningeal hemorrhages, leading to hemosiderin deposition along the superficial cortical layers.^12^ These bleeds may result from the rupture of fragile, amyloid-laden pial vessels. Individuals with CAA-related cSS often show rapid disease progression and are at increased risk for future fatal ICH.^13, 14^ Preliminary evidence, including from our cohort, suggests that corticosteroid therapy induces regression of vessel wall enhancement and inflammation.^4, 15^ Whether meningovascular inflammation contributes to vessel wall fragility and bleeding susceptibility, and whether anti-inflammatory strategies can modify long-term outcomes, including survival and hemorrhage risk, warrants further investigation.

### Limitations

This study is limited by its retrospective design, small sample size and lack of pathological confirmation of vascular inflammation. These factors restrict generalizability and introduce potential selection bias. Nonetheless, the consistency of neuroimaging findings and supportive CSF data offer compelling indirect evidence for a clinically relevant meningovascular inflammatory process. Time-of-flight MR angiography was not routinely acquired, limiting systematic correlation of VWI findings with vascular anatomy. While slow-flowing veins can appear hyperintense, we aimed to exclude venous structures based on pre-contrast sequences and typical morphology. Although contrast-enhanced FLAIR sequences were not systematically acquired, they have also shown utility in detecting leptomeningeal enhancement in CAA and could be a practical alternative in settings with limited access to VWI protocols.^16, 17^ It may be particularly useful on lower field-strength scanners. However, unlike VWI, it does not permit direct visualization of mural vessel enhancement. Future prospective studies with larger cohorts are warranted to validate these observations and evaluate the effect of anti-inflammatory treatment on clinical outcomes in patients with CAA-related cSS.

## Data Availability

All data produced in the present study are available upon reasonable request to the authors.

## Acknowledgments

This study was supported by the BB-DARS project, funded by the Deutsche Alzheimer Gesellschaft (DAlzG) and the Förderstiftung Dierichs.

## Author Contributions

S.S. contributed to the conception and design of the study; all the authors contributed to the acquisition and analysis of data; P.A., S.S. and A.C. contributed to drafting the text; P.A., E.K. and S.M. contributed to preparing the figures. All the authors contributed to a critical review of the manuscript.

## Data Availability

The corresponding author has full access to the data used in this manuscript and all data are available on reasonable request.

